# The use of simultaneous SEEG and MEG in localising seizure onset zone

**DOI:** 10.1101/2020.01.31.20019505

**Authors:** Umesh Vivekananda, Chunyan Cao, Wei Liu, Jing Zhang, Fergus Rugg-Gunn, Matthew C Walker, Vladimir Litvak, Bomin Sun, Shikun Zhan

## Abstract

**Objectives:** Both Magnetoencephalography (MEG) and stereo-electroencephalography (SEEG) are used in presurgical epilepsy assessment, with contrasting advantages and limitations. It is not known whether combined recording using both modalities can maintain inherent advantages whilst overcoming these limitations e.g. recording from deep brain sources whilst preserving good spatial resolution.

**Methods:** 24 adult and paediatric patients who underwent SEEG study for pre-surgical evaluation of focal drug-resistant epilepsy, were recorded using simultaneous SEEG-MEG, of which 14 had abnormal interictal activity during recording. The 14 patients were divided into two groups; those with presumed superficial (n=7) and deep (n=7) brain interictal activity.

**Results:** There was no significant difference between SEEG and MEG in identifying superficial spikes (p=0.135) and SEEG was significantly better at detecting deep spikes (p=0.002). Mean distance across patients between SEEG channel with highest average spike amplitude and MEG dipole was 26.6+/-3.6 mm for superficial sources, and 21.5 +/- 2.04 mm for deep sources, even though for some of the latter (n=4) no MEG spikes were detected and MEG dipole was fitted to a SEEG interictal activity triggered average. Removal of MEG dipole was associated with 1 year seizure freedom in 5/6 patients with superficial source, and 4/6 patients with deep source.

**Conclusions:** Although SEEG has greater sensitivity in identifying interictal activity from deeper sources, an MEG source can be localised using SEEG information, thereby providing useful whole brain context to SEEG and potential role in epilepsy surgery planning.

## Introduction

Magnetoencephalography (MEG) and stereo-electroencephalography (SEEG) can provide complementary information for presurgical epilepsy assessment. MEG is non-invasive and has high temporal and spatial resolution with good global coverage ^1,2^. However, deep sources, such as the mesial temporal lobe, a region commonly associated with refractory epilepsy, are poorly detected with MEG ^3^. SEEG is an invasive procedure, in which a limited set of electrodes are placed within the brain; these provide excellent detection of nearby sources but have restricted spatial sampling. Thus, a method to combine both modalities would be advantageous, with MEG providing whole brain information, and SEEG providing information on deep sources.

MEG has previously been compared with SEEG non-concurrently ^4,5^. Few studies have reported simultaneous SEEG and MEG recordings, most likely due to technical challenges in its acquisition ^6,7^. Here we report a case series of simultaneous SEEG and MEG recordings in a range of refractory epilepsies, and directly compare interictal epileptiform activity (IED) and its localisation. In addition, we relate these findings to the outcome in patients who underwent epilepsy surgery.

## Methods

### Simultaneous SEEG-MEG recordings

The study included 24 patients (adults and children) who underwent SEEG study for pre-surgical evaluation of focal drug-resistant epilepsy at Shanghai Jiaotong University School of Medicine. The study was approved by the local ethics committee of Affiliated Ruijin Hospital, Shanghai Jiaotong University School of Medicine and written consent was obtained from patients or carers. Location and number of SEEG electrodes implanted varied between patients depending on presumed epileptogenic focus (Table e-1).

Simultaneous ∼7 minute SEEG-MEG recordings were performed with a 306-channel, whole-head VectorView MEG system (Elekta Oy, Helsinki, Finland) in a magnetically shielded room (Euroshield, Eura, Finland). The raw MEG data were band pass filtered 0.03-330 Hz and digitized at 1000 Hz. The magnetic artefacts and movement artefact were removed by the temporal extension of Signal Space Separation method (tSSS) implemented in the MaxFilter software (Neuromag 3.4, Elekta Oy, Helsinki, Finland). 10 patients had no epileptiform activity on SEEG and so were excluded from further analysis. One patient had a seizure.

### SEEG analysis

Analysis was performed using Brain Electrical Source Analysis software (BESA GmbH, Germany, http://www.besa.de/), Statistical Parametric Mapping (SPM12, UCL, https://www.fil.ion.ucl.ac.uk/spm/), and Fieldtrip (http://fieldtriptoolbox.org). SEEG was analysed using bipolar montage (Band-pass filter 1Hz-70Hz, 50Hz notch). Interictal spikes were identified using BESA software. All spikes were manually marked at the peak of maximal positive/negative deflection of the spike. The electrode contact with the largest average spike amplitude was then noted (annotated as ‘best’ channel).

Locations of implanted SEEG electrodes were identified from postoperative CT scans using Lead-DBS toolbox (https://www.lead-dbs.org/). Post-operative CT was co-registered with a pre-operative T1 structural MRI in SPM12 and further adjusted under manual control using Slicer software (https://www.slicer.org/). SEEG contact locations were then obtained by manually fitting electrode models to the artefacts seen in the CT using the interface implemented in Lead-DBS.

### MEG analysis

Analysis of the MEG recording was performed ‘blind’ to SEEG findings. Interictal spikes were identified using BESA software (Band-pass filter 1-35 Hz, 50Hz notch, gain 400-800fT). All spikes were manually marked at the peak of maximal positive deflection of the spike.

Source localisation for MEG data was performed by averaging individual spikes (BESA software), before importing into SPM. A time window was then set 100ms before the rise phase and 100ms after the fall phase of the average spike. A single shell forward model ^8^ based on canonical meshes inverse normalised ^9^ to the pre-operative T1 structural MRI image was used to fit a single dipole (Fieldtrip) and the corresponding residual variance image was also examined. If there were no spikes seen on MEG alone, MEG source activity (M-source) was derived by averaging the raw MEG data informed by co-existent SEEG spikes (taken 1 second before and after highest amplitude of spike), before the same dipole fitting process was performed.

Where possible, post-resection MRI images were co-registered to the pre-operative T1 structural MRI to assess post-surgical outcome.

### Data Availability Statement

Anonymized data will be shared by request from any qualified investigator

## Results

### 1. SEEG and MEG are equally sensitive in identifying superficial interictal activity but not deep brain interictal activity

For patients whose presumed epileptogenic focus was superficial cortex, as defined by SEEG best channel, we found no statistical difference between number of spikes identified by SEEG versus MEG (paired t-test, p=0.135) (Figure 1A). This suggests that SEEG and MEG are equally sensitive in identifying interictal spikes from cortical sources. For patients with a presumed deep brain epileptogenic focus, we found that number of spikes identified by SEEG was significantly higher than MEG (Mann Whitney U test, T=56.5, p=0.002), indicating that SEEG is more sensitive than MEG in identifying interictal spikes from deep sources (Table 1).

**Table 1.** Number of interictal spikes identified on SEEG alone, SEEG and MEG, and MEG alone for presumed superficial and deep brain sources. ‘Best’ channel indicates SEEG channel with highest amplitude average spike.

| Source | Region | Patient | SEEG alone | SEEG + MEG | MEG alone | 'Best' channel |
| --- | --- | --- | --- | --- | --- | --- |
| Superficial | Frontal | 2 | 5 | 18 | 3 | Right F 6-7 |
|  | Temporal | 3 | 10 | 6 | 0 | Left mesial T 6-7 |
|  |  | 4 | 4 | 11 | 4 | Right HPC 6-7 |
|  |  | 5 | 1 | 14 | 0 | Left T 4-5 |
|  |  | 10 | 5 | 17 | 3 | Right T 8-9 |
|  |  | 11 | 0 | 4 | 2 | Left T 9-10 |
|  | Parietal | 6 | 0 | 21 | 0 | Left P 6-7 |
| Deep | Temporal | 1 | 22 | 1 | 0 | Left HPC 3-4 |
|  |  | 7 | 26 | 0 | 0 | Right T I 0-1 |
|  |  | 9 | 7 | 4 | 1 | Left HPC 0-1 |
|  |  | 12 | 34 | 0 | 0 | Left HPC 0-1 |
|  |  | 13 | 26 | 0 | 0 | Left HPC 3-4 |
|  |  | 14 | 30 | 0 | 0 | Right HPC 2-3 |
|  | Parietal | 8 | 4 | 1 | 0 | Left P 1-2 |

**Figure 1.**
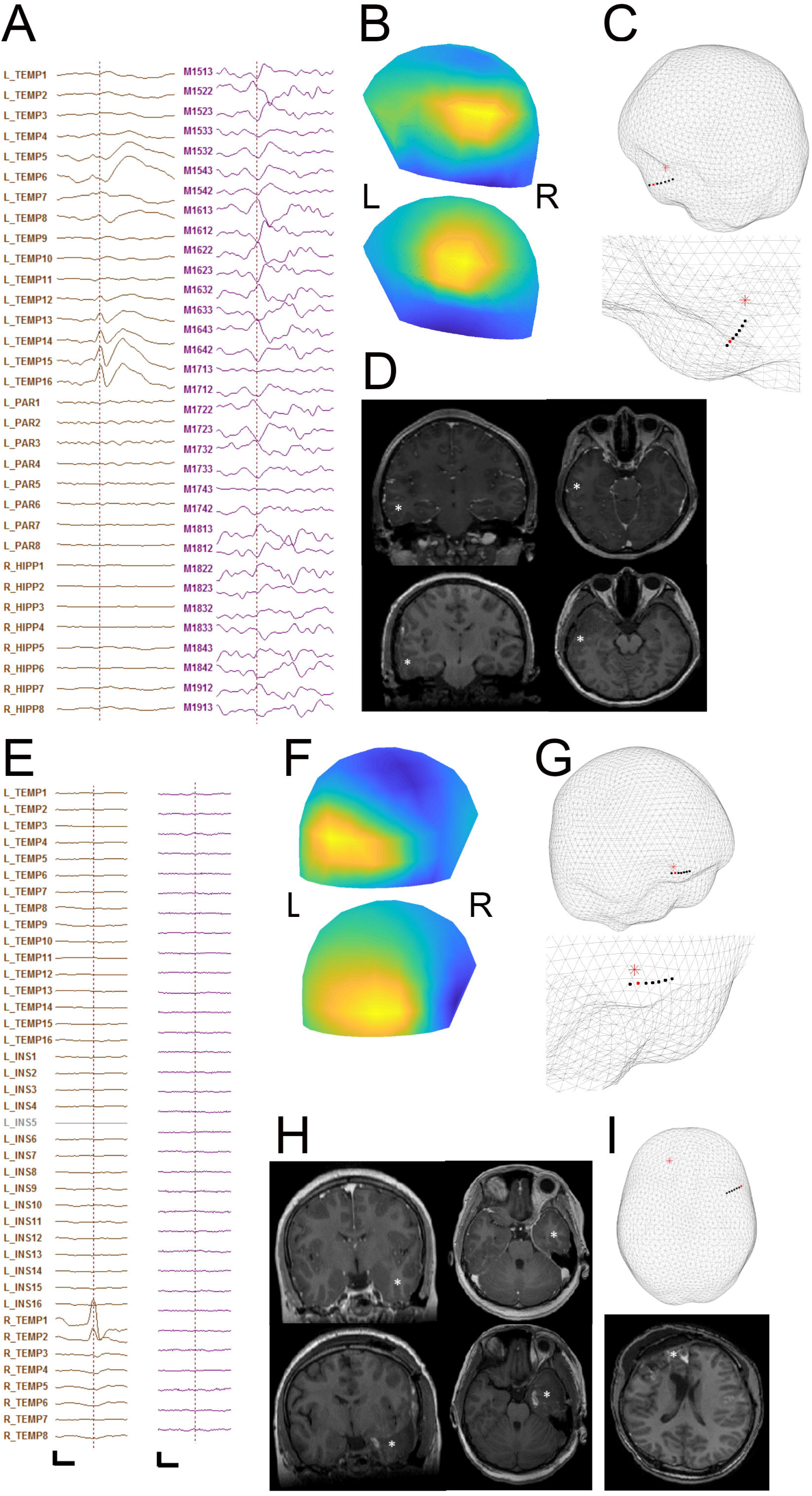
A. Example of superficial region interictal spike from SEEG (left) and MEG (right) B. Field map (measured top and modelled bottom) corresponding to the peak of the average epileptic spike, L=left and R=right C. Relation on inner skull mesh of MEG dipole (red star) and electrode contact (red circle) with highest spike amplitude, top, and zoomed region, below D. Upper panel is the position of MEG dipole (white asterix) on pre-operative MRI scan coronal and sagittal planes; lower panel is post-operative MRI scan. E. Example of deep region interictal spike from SEEG (left) and MEG (right). Scale bar: 300μV/500fT and 0.2s. F. Field map corresponding to the peak of the average epileptic spike G. Relation of MEG dipole and electrode contact with highest spike amplitude H. Position of MEG dipole on pre-operative MRI (upper) and post-operative MRI (lower) I. Difference in interictal source localisation between MEG dipole (red star) and SEEG best channel (red circle), top, and MEG dipole on post-operative scan for that patient, bottom.

### 2. Averaged MEG spike source localisation is broadly concordant with SEEG localisation for superficial epileptogenic regions, and relates well with post-operative outcome

We examined the relationship of MEG source activity location, and location of SEEG best channel. For patients with presumed superficial epileptogenic regions, the mean distance between M-source and ‘best’ channel was 26.6+/- 3.6 mm., suggesting that location of average MEG source activity is closely related to average SEEG interictal spike location for superficial epileptogenic regions (Figure 1C). In two patients (4 and 11), although SEEG identified interictal activity in the lateral temporal lobe during simultaneous recording, subsequent SEEG assessment of seizures located the seizure onset zone to be in frontal and occipital brain regions respectively. However, unlike SEEG, MEG during simultaneous recording accurately source localised to these regions (Figure 1I).

Success of MEG dipole removal during epilepsy surgery was then related to seizure recurrence after 12 months. In all patients MEG dipole location was concordant with presumed epileptogenic zone and removed brain region (Table 2), confirmed by post-operative MRI in 3 patients (Figure 1D). In five out of six cases (except patient 6) there was no seizure recurrence.

**Table 2.**
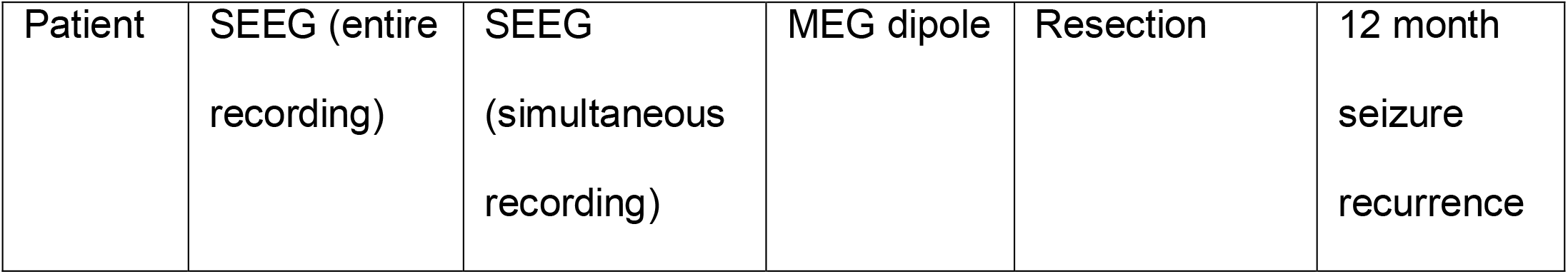
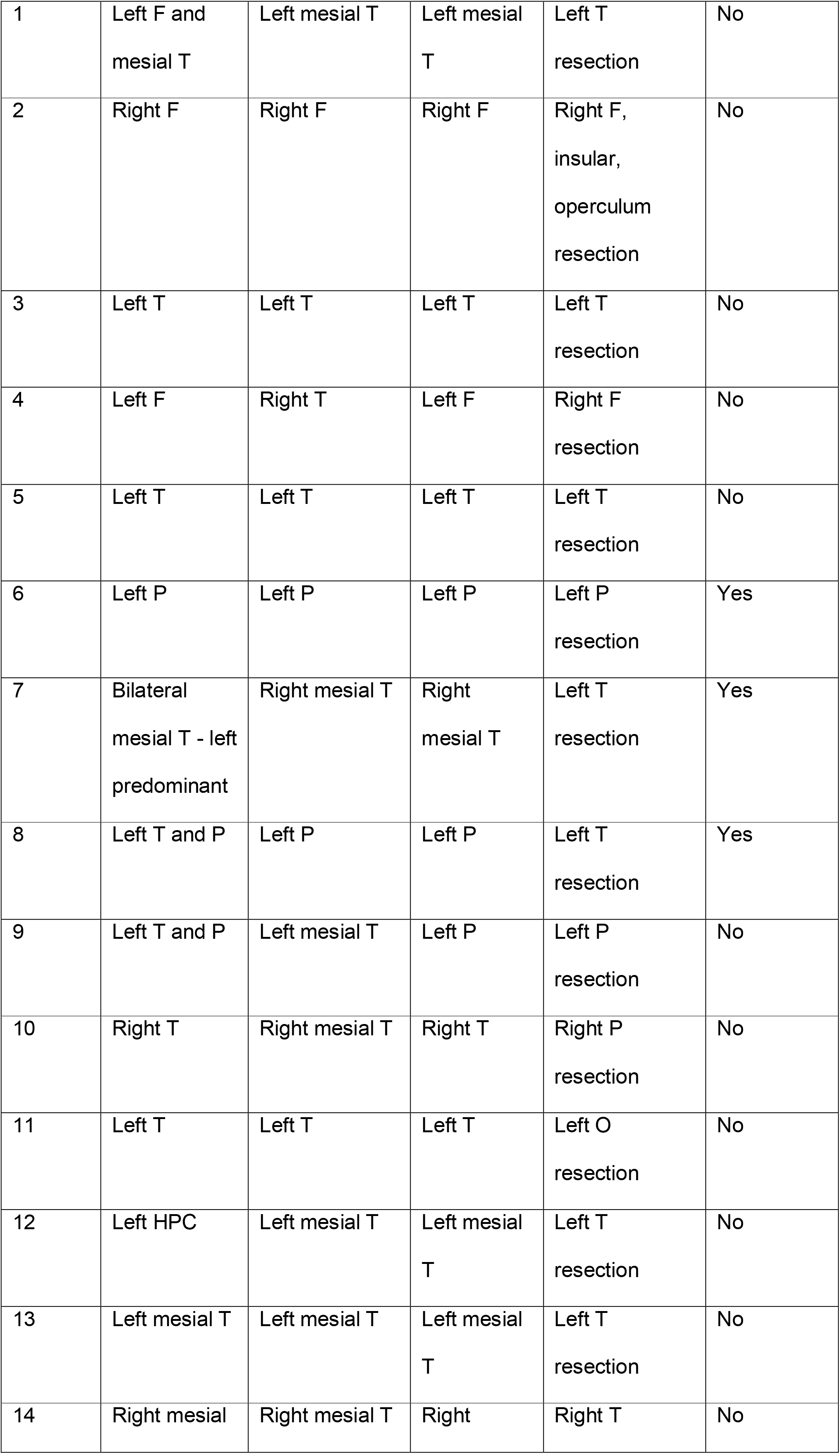
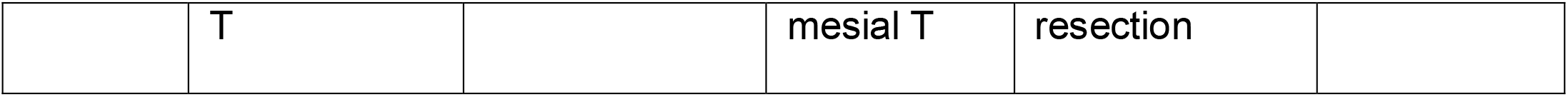
Post surgery outcomes. SEEG entire recording describes location derived from prolonged telemetry and SEEG simultaneous recording describes location derived from simultaneous SEEG/MEG recording. F=frontal, T=temporal, P=parietal, O=occipital, HPC=hippocampus

### 3. MEG localisation of interictal activity for deep sources can be informed by SEEG, and relates well with post-operative outcome

For patients with presumed deep epileptogenic regions, the mean distance between M-source and ‘best’ channel was 21.5 +/- 2.04 mm. In four mesial temporal cases no MEG spikes were identified, meaning that M-source was informed from SEEG spikes instead (Figure 1E). Interestingly even in these cases location of M-source was closely related to ‘best’ channel location (Figure 1G), suggesting that simultaneous MEG and SEEG have complementary localising value, even in cases where no apparent MEG interictal activity is seen.

In five out of six patients MEG dipole location was concordant with presumed epileptogenic zone and the removed brain region (Table 2), confirmed by post-operative MRI in 3 patients (Figure 1H). In four out of six patients (except Patients 7 and 8) there was no seizure recurrence. The MEG dipole location in patient 8 (parietal lobe) was presumably outside of the resected brain region (temporal lobe), possibly explaining persistence of seizures.

## Discussion

In this large study of epilepsy patients undergoing simultaneous SEEG-MEG study, we could directly compare the sensitivity for both modalities in identifying interictal spikes and spike localisation. As in previous studies we found that SEEG and MEG were comparable in identifying interictal spikes originating from superficial cortex, with MEG identifying spikes not viewed on SEEG in a number of patients. This likely reflects the relative limited spatial sampling SEEG provides. In contrast, SEEG was far superior in identifying spikes originating from key deep brain regions (e.g. mesial temporal). This has generally been perceived as a weakness of MEG both in clinical evaluation and in normal physiological studies.

Surprisingly, although MEG did not identify any interictal spikes for the majority of patients with deep sources, average MEG activity informed by identified SEEG spikes still accurately localised deep source activity. This is in keeping with the recent observation that SEEG informed deep brain MEG activity can be detected using independent component analysis (ICA) ^10^. Here we further show that surgical resection of the consequent average MEG dipole predicted seizure freedom (5 out of 6 patients); the one patient where the dipole was not removed had seizure recurrence.

There were limitations of the study, including the brevity of recordings, which was dictated by the technical difficulty of acquiring simultaneous SEEG and MEG data. Secondly, the implantation plan was variable between patients, and therefore direct comparison of SEEG and MEG needs to be examined with this in mind.

Simultaneous SEEG and MEG can thus provide complementary information about the spatial extent of interictal epileptiform activity for superficial and deep epileptogenic sources, and so better inform surgical planning.

## Data Availability

Anonymized data will be shared by request from any qualified investigat

## Acknowledgements

We would like to thank all the patients who took part in this study. We would like to thank Andreas Horn and Ningfei Li for their help with adding support for iEEG electrode reconstruction to Lead-DBS.

